# Correlations between patterns of activity and the response to treatment yield distinct signatures for different antidepressive treatments

**DOI:** 10.1101/2023.09.29.23294935

**Authors:** Stefan Spulber, Frederik Elberling, Sandra Ceccatelli, Martin Gärde, Mikael Tiger, Johan Lundberg

**Affiliations:** Department of Neuroscience, Karolinska Institutet, Stockholm, Sweden; Centre for Psychiatry Research, Department of Clinical Neuroscience, Karolinska Institutet & Stockholm Health Care Services, Region Stockholm, Karolinska University Hospital, Stockholm, Sweden

## Abstract

Alterations in circadian patterns of activity have been described in several psychiatric disorders, and in major depressive disorder (MDD) have been associated with symptom severity. The aim of this study was to investigate the correlations between activity patterns and the response to different antidepressive treatments. We used actigraphy recordings acquired in three independent clinical trials investigating the response to internet-delivered cognitive behavioral therapy (iCBT), escitalopram, or ketamine, where symptom severity was assessed on MADRS scale. Data processing procedure was designed to extract features describing individual circadian cycles, as well as the average circadian profile for each subject. The analysis of effects of antidepressive treatments on single features (independently from magnitude of response) showed alterations in treatment-specific subsets of features. Further analysis revealed distinct correlations between longitudinal changes in individual features and the response to iCBT, escitalopram, or ketamine. Linear regression ensembles modeling the response to treatment were trained on actigraphy recorded during the depressive episode, before intervention. The analysis of coefficients following Bayesian model averaging showed that better response to iCBT was associated with weaker circadian entrainment; better response to ketamine was associated with more robust circadian entrainment and higher fragmentation of activity patterns; while higher age and steeper propensity to sustain activity was associated with better response to either iCBT or ketamine. Our data suggest that the analysis of circadian patterns of activity can potentially be used for predicting the response to specific MDD treatments, however larger confirmatory studies are required.

**Highlights:**

- Different antidepressant treatments alter specific subsets of features of activity patterns.
- The magnitude of response to treatment correlate with changes in patterns of activity.
- Patterns of activity correlate with the response to specific antidepressive treatments.

## Introduction

Major depressive disorder (MDD) is a highly prevalent mental disorder with heterogenous biological background, and there are many antidepressive interventions available (Khan et al., 2012). Cognitive behavioral therapy (CBT) is well-established antidepressive non-pharmacological intervention (Weitz et al., 2015). Internet-delivered CBT (iCBT) has been shown to be as effective as face-to-face and has the advantage of using standardized treatment modules to ensure homogenous intervention suitable for longitudinal studies (Carlbring et al., 2017). Antidepressant drugs act on specific neurotransmitter signaling pathways, including serotonin, dopamine, noradrenaline, and glutamate. Selective serotonin reuptake inhibitors (SSRI) are the most commonly prescribed antidepressants, and it typically takes several weeks for the antidepressive effects to stabilize (Machado-Vieira et al., 2010). The introduction of rapid acting antidepressants, such as ketamine in subanesthetic dose (Berman et al., 2000) lead to a paradigm shift in the treatment of depression (Krystal et al., 2019).

The prediction of response to treatment is relevant particularly for the wellbeing of the patient as well as for avoiding unnecessary consumption of healthcare services. Several attempts have been described in the literature, including the change in reaction to facial expression early after starting the treatment (Browning et al., 2019), based on the patterns of change in scores for individual items after 4 weeks of treatment (Athreya et al., 2021), or using combinations of genetic and non-genetic biomarkers (Taliaz et al., 2021). While carrying undeniable advantages towards personalizing the MDD treatment, they either rely on treatment to be initiated before evaluation or require collecting biological samples for genotyping. Recording of movement by means of wrist actigraphy provides a promising non-invasive technology for data collection, and accumulating evidence points to associations between specific alterations in activity and different psychiatric disorders (Chapman et al., 2017; Kang et al., 2024; Pan et al., 2014; Tazawa et al., 2019). For MDD, the most common alterations reported are lower total amount of activity and blunted circadian rhythms (Lyall et al., 2018; Morres et al., 2019a). Similar associations have been described between activity levels and severity of depressive symptoms (Helgadóttir et al., 2015; Morres et al., 2019b), and we have shown recently that the patterns of activity correlate with depression severity independent of actual levels of activity (Spulber et al., 2022). However, the potential correlations between activity patterns and the response to treatment have received less attention to date.

The aim of this study was to investigate the correlations between activity patterns and the response to different MDD treatment alternatives. To this end we designed a features extraction procedure including both sequence- and circadian profile-based features. We first explored the effects of different antidepressive treatments on circadian patterns of activity in MDD subjects. Second, we explored the changes in patterns of activity in relation to the magnitude of response to treatment. Lastly, we trained multivariate linear regression ensembles to fit the response to treatment and analyzed the impact and implications of individual features.

## Materials and methods

### Data collection

We pooled together actigraphy recordings from 3 independent studies: (1) a study on serotonin transporter availability in patients given internet-based Cognitive Behavioral Therapy (iCBT) for the treatment of a major depressive episode (Svensson et al., 2021) (Ethical Permit No. 2014/452-31, 2015/1177-32, Swedish Ethical Review Authority; pre-registration: AsPredicted.org #14118). Healthy controls (HC, N=17) and subjects suffering from depression (N=17) were recruited by advertisements in local newspapers. Diagnosis of depression was established after full psychiatric assessment by a psychiatrist or by a resident physician supervised by a senior psychiatrist using the Mini International Neuropsychiatric Interview (M.I.N.I.). The study included patients with an ongoing major depressive episode according to DSM-IV criteria, with a history of at least one prior episode, and which were not undergoing any psychopharmacological treatment for MDD. Eligible patients had a MADRS score between 18 and 35. HC had no history of psychiatric illness and matched the patients by age, sex, and intellectual ability. The iCBT protocol consisted of 10 modules delivered in an online program. Throughout the program, the patients had been assigned to a psychologist who supervises the progress and provides individual feedback (Svensson et al., 2021). The study participants were instructed to wear the actigraph (GENEActiv, ActivInsights, Cambs, UK) continuously on the wrist of the non-dominant arm. In MDD patients, actigraphy recordings were acquired before starting, and after completing the iCBT program. (2) a study on the effects of escitalopram treatment on serotonin 1B receptor binding in MDD (Gärde et al., 2024) (Ethical Permit No. 2018/1403-31/1, Swedish Ethical Review Authority; pre-registration: AsPredicted.org #33267). The patients were recruited via online advertising and from Gustavsberg University Primary Care Center (Stockholm County, Sweden). The MDD diagnosis was confirmed by a psychiatrist according to M.I.N.I. Patients with MADRS>20 without ongoing antidepressant treatment and medical history free from CNS or somatic diseases (N=8) received escitalopram 10mg/day for 8 weeks. MADRS was recorded before treatment initiation, and treatment response was evaluated after 3-4 and 6-7 weeks treatment with escitalopram (Gärde et al., 2024). Actigraphy recordings were acquired using actigraphs (Actiwatch 2, Philips Respironics, Murrysville, PA, USA) worn on the wrist of the non-dominant arm for ∼1 week before treatment initiation, and after 3 weeks of escitalopram treatment. (3) a study on the effects of ketamine on serotonin 1B receptor binding in patients with selective serotonin reuptake inhibitors (SSRI) resistant depression (Tiger et al., 2020) (Ethical Permit No. 2017/799-31, Swedish Ethical Review Authority; pre-registration: AsPredicted.org #17602). The study recruited by internet-based advertisements, and included patients with an ongoing major depressive episode (according to M.I.N.I.), with MADRS ≥ 20, resistant to SSRI treatment in an adequate dose for at least 4 weeks. Ongoing antidepressant treatment was discontinued and actigraphy data was collected after a washout period of at least 5 times the half-life of the SSRI. The patients were instructed to wear the actigraph (Actiwatch 2, Philips Respironics, Murrysville, PA, USA) continuously on the wrist of the non-dominant arm and not remove it unless for personal safety reasons. The recording started prior to the first ketamine or placebo infusion and continued for the duration of the open-label ketamine treatment program (0.5mg/kg, 2 infusions/week for 2 weeks; placebo N=10, ketamine N=20) (Tiger et al., 2020). Basic demographics, including the number of subjects included from each study are depicted in Table 1. Preliminary analyses did not identify significant differences between male and female subjects in individual features, symptom severity, or response to MDD treatment, therefore sex was not included in further analyses. The data available in the three cohorts allowed alternative partitioning into “before treatment” (MDD, pooled N=35), which was used as reference for all subsequent comparisons involving subjects on antidepressive treatments. Similarly, all subjects with MADRSpost ≤ 10 were assigned to a single group “remission” (pooled N=12, including iCBT, N=3; escitalopram, N=2; ketamine, N=7).

**Table 1.** Description of patient cohorts and antidepressant treatments.

| Treatment | pre | post | both<sup>1</sup> | Treatment scheme | Ref |
| --- | --- | --- | --- | --- | --- |
| HC | 19<sup>2</sup> (14) | N/A | N/A | N/A | (Svensson et al., 2021) |
| iCBT | 12 (9) | 11 (9) | 8 (6) | 10 self-administered, internet-delivered sessions (1 session/week) | (Svensson et al., 2021) |
| Escitalopram | 7 (3) | 6 (2) | 5 (2) | 10 mg/ day | (Gärde et al., 2024) |
| Ketamine | 16 (8) | 15 (6) | 12 (6) | 0.5 mg/kg i.v. infusion, 2/week | (Tiger et al., 2020) |

Numbers in parentheses indicate the number of female subjects in each group.
<sup>1</sup> – used for paired comparisons, post vs. pre
<sup>2</sup> – 1 subject was recorded 3 times; 1 subject was recorded 2 times; 16 independent subjects

The timing of actigraphy recording and MADRS evaluations are depicted in Fig. 1A, and the number of days of actigraphic recordings are depicted in Supplementary Table 1. Actigraphic recordings on patients receiving iCBT as MDD treatment were acquired using GENEActiv Original wrist-worn actigraphs (ActivInsights, Cambs, UK). The devices use three-dimensional accelerometers (dynamic range up to 8*g*; 12-bit encoding, resolution 3.9 m*g*) at 30 Hz sampling rate to record wrist movement. The raw data was downloaded using proprietary software, then processed in MATLAB (The Mathworks, Natick, MD, USA), using a modified version of the code (https://github.com/DavidRConnell/geneactivReader), as described earlier (Ekholm et al., 2020). Briefly, the Euclidean norm of change in acceleration vector was first smoothed using a rolling Gauss window spanning 30 consecutive datapoints (1s), then a high-pass filter was applied (threshold: 20 m*g* = 196 mm/s^2^) before computing the sum of changes in acceleration vectors over 1 min epochs (1440 samples/24h). Actigraphic recordings on patients receiving escitalopram or ketamine as antidepressant treatment were acquired using Actiwatch 2 wrist-worn devices (Philips Respironics, Murrysville, PA, USA) set to record activity only integrated over 1 min epochs. The raw data was downloaded according to manufacturer’s instructions (Actiware 6.x, Philips Respironics) then exported as text files. The text file was imported to MATLAB using a custom function designed to yield an output similar to the one generated by the import function for GENEActiv devices.

**Fig. 1.**
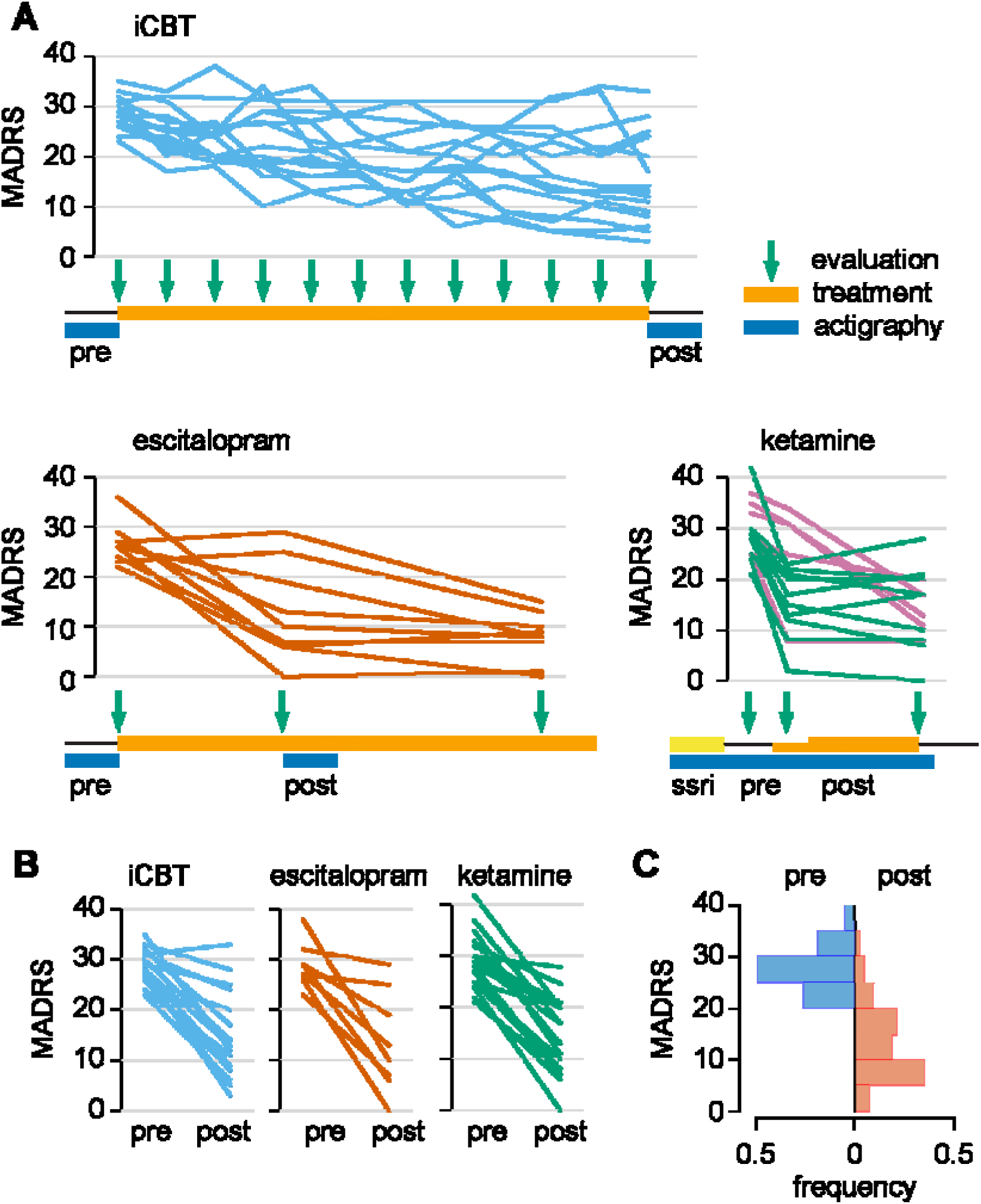
Description of source data. (**A**) Timeline for data collection for all cohorts included. Only subjects for which actigraphy recordings were available are shown. (**B**) Change in MADRS score in individual subjects for each cohort. (**C**) Distribution of MADRS scores before and after treatment in pooled cohorts. The subjects with MADRSpost < 10 are included in the “remission” group in subsequent analyses.

### Quality control and inclusion criteria

The quality control was performed by the same observer, blind to group belonging. All recordings were first inspected visually using a standardized procedure designed to identify stretches of missing data, artifacts, and gross abnormal circadian patterns of activity (*e.g.*, shift work, or other consistent activity at night). Intervals containing suspected shift work (not reported at the time of recording), potential artifacts, or missing data, were cropped out. Only recordings spanning at least 3 consecutive days were included in further analyses.

### Pre-processing and feature extraction

All processing of actigraphy data was performed in MATLAB. The data import procedures yielded sizeable differences in range of raw output (activity/min) between GENEActiv and Actiwatch 2 devices, but the coefficient of variability of individual days was similar across devices (see Supplementary Fig. S1). These differences were managed in the design of feature extraction procedures as follows: (1) no features relying on absolute magnitude values were included; and (2) features such as circadian peak and trough were calculated after mean normalization of raw data. We first cropped all recordings between the first and the last recorded midnight to yield an integer number of 24-h periods. For each subject we applied the feature extraction procedures on individual days and on circadian profiles calculated as the average across corresponding time bins of registered individual days. The features calculated on individual days were used for estimation of variability across days.

The features we extracted describe the regularity, fragmentation, and complexity of circadian patterns of activity, and uses custom implementations of publicly available algorithms. The complete list of features and their calculations is available in Supplementary Table 2. The following features were extracted: circadian period; circadian peak and trough; relative amplitude (Ekholm et al., 2020; Gonçalves et al., 2015; Lyall et al., 2018); scaling exponents (Hu et al., 2004); intradaily variability (IV); interdaily stability (IS) (Gonçalves et al., 2014); day-to-day variability, and propensity to sustain activity.

Circadian period was estimated using the Lomb-Scargle algorithm optimized for MATLAB implementation (Saragiotis, 2021). The Lomb-Scargle periodogram was preferred over the commonly used Sokolove-Bushell algorithm (Sokolove and Bushell, 1978) because the latter has been shown to yield period estimates biased towards periods below 24 h (Tackenberg and Hughey, 2021). The circadian period was calculated over the entire recording using an oversampling factor of 10 to yield resolution of the estimated in the range of minutes. The scaling exponent for detrended fluctuation analysis was calculated for the magnitude of measured activity in 1-min bins using boxes equally spaced on a logarithmic scale between 4 min (4 consecutive samples) and 24 h (1440 consecutive samples) as described by Hu et al. (Hu et al., 2004). The scaling exponent is a feature of the intrinsic regulatory mechanisms controlling the rest/activity patterns. It has not been shown to be sensitive to extrinsic factors the subject is exposed to in normal daily activity, but is altered as a result of disease (Chapman et al., 2017; Fasmer et al., 2016; Hu et al., 2004). Intradaily variability (IV) estimates the fragmentation of activity patterns by calculating the ratio between mean squared differences between consecutive time intervals and the mean squared difference from global mean activity per interval; it increases as the frequency and the magnitude of transitions between rest and active intervals increase, and decreases as the active and inactive intervals consolidate (Gonçalves et al., 2015). Interdaily stability (IS) evaluates the coupling between activity patterns and circadian entrainers and is calculated as the ratio between variability around circadian profile and global variability. High values indicate consistent activity patterns across days, consistent with strong coupling between activity and circadian entrainers. The relative amplitude of circadian rhythms of activity (RA) estimates the robustness of average circadian rhythms (Edgar and McClung, 2013; Lyall et al., 2018). The range of RA is bounded between 0 (no circadian rhythms) and 1 (robust circadian rhythms, with consistent timing of consolidated rest interval longer than 5 h across days). The day-to-day variability comprised 3 features as follows: circadian profile variance between consecutive days (SeqVar), calculated as Euclidean distance between consecutive days, normalized to the total number of samples per day; variation from average circadian profile (ProfileVar), calculated as the Euclidean distance between each day and the average profile, normalized to the total number of samples per day; and the normalized difference between consecutive days (SeqProfileVar), calculated as the ratio between mean difference between circadian profiles of consecutive days and mean deviation from average circadian profile. The propensity to sustain activity (SustainProp) was calculated as the slope of likelihood to sustain or increase activity in the next minute against current level of activity. The distribution was calculated for minutes with activity count>10 (which eliminated the range specific for sleep, ∼30% of datapoints/day) in 20 equally spaced bins covering the range up to the 99^th^ percentile of active minutes. This is a hybrid measure applying a sequence-based analysis on the distribution of activity counts/min (assumed to have exponential distribution). The likelihood to further increase activity drops with increasing the activity counts in the current minute, therefore the slope is negative, and approaches 0 at the right tail of the distribution. A shallow slope indicates the subject is unlikely (not willing) to sustain even low levels of activity. For scrambled data (preserved distribution, but random sequence), the slope is about -0.3 (likelihood to sustain activity decreases by 30% when the activity count increases 10-fold).

### Feature selection

There is virtually no consensus around the analysis of actigraphy and parameter selection for feature extraction. Therefore, the initial feature space included a number of features with different degrees of similarity, as illustrated by the matrix of correlations in Supplementary Fig. S2. To mitigate the impact of multicollinearity, one can choose between dimensionality reduction techniques (*e.g.*, PCA, UMAP), and feature selection. The former has the advantage of identifying latent structures in the feature space and accurately accounting for underlying patterns in downstream calculations, but the analysis of individual feature contribution is not straightforward. The latter facilitates the intuitive interpretation of the final result, but carries the risk of missing out on underlying latent structures. We opted for feature selection to take advantage of the interpretability of the results. This is particularly relevant for Bayesian model averaging, where posterior inclusion probability is artificially deflated by including highly correlated features. The initial feature space included 90 features, distributed in 4 overlapping clusters as follows: circadian profiles; mean daily features; daily variability around the average; and sequential daily variability. Day-to-day variability (calculated as the ratio between mean squared sequential differences in circadian profile and the mean squared difference from average profile) could only be calculated on the full recording and was assigned to the first cluster. We used a hybrid (heuristic and data driven) approach for feature selection, focusing on reducing the clusters of conceptually related features which were highly correlated in healthy controls and MDD patients before treatment (see Supplementary Fig. S3). The correlation patterns yielded by features calculated on circadian profiles largely matched the correlations among features calculated on single days. In addition, the correlations between variability around the average and sequential variability were virtually perfect, rendering the 2 sets redundant. The final feature space consisted of 28 features, including 12 from the circadian profile cluster and their correspondents in daily variability around the average; propensity to sustain activity and its variance across days; day-to-day variability of circadian profile; and subject age (Supplementary Table 2).

### Effects of antidepressive treatments and correlations with response to treatment

The design of analysis uses several underlying assumptions. (1) The effect of antidepressant treatment is assumed to have reached steady state at the time of recording, and no features exhibit significant drifting. This was verified by inspection of trends in individual features for each subject (not shown). (2) MADRS evaluation reflects an average state spanning the best part of the recording period. The self-assessment MADRS scale requires the subject to evaluate the severity of symptoms over the last 3 days, and it is therefore justifiable to assume irrelevant changes over a period of 5-7 days around the time of evaluation. (3) The pharmacological effects are stable on both mood and circadian regulation of activity. Classical antidepressive treatments take a couple of weeks to exert significant effects on mood (*i.e.*, iCBT and escitalopram), which is then stable for sufficiently long time for the purpose of actigraphy recording. Fast-acting antidepressants (*i.e.*, ketamine), induce measurable effects on mood within a couple of hours after administration. The regimen of ketamine administration in the cohort analyzed here was designed to ensure stable drug effect covering 2 weeks in the open-label phase of the study.

The response to treatment was calculated as relative change from baseline MADRS. The MADRS scores used for the estimation of response to treatment were the last measurement available before starting the treatment (baseline), and the last measurement available after treatment initiation (see also Fig. 1C, D for details).

### Ensemble training and Bayesian model averaging

We implemented an ensemble approach to fit the response to treatment, as described earlier (Spulber et al., 2025). The training was performed independently for 3 groups: iCBT, ketamine, and any treatment (iCBT, ketamine, or escitalopram). We have generated the initial ensemble by independent homogenous training using a systematic bootstrapping (with replacement) scheme for selecting up to 6 features/model (systematic testing of all possible combinations of features). This approach ensured a minimum of 2 subjects/feature for the iCBT cohort (N=12). The number of subjects available in the escitalopram cohort (N=7) was too low to train meaningful models, therefore the response to escitalopram was not modeled independently. After training, we applied an adaptation of Occam’s window algorithm (Madigan and Raftery, 1994; Raftery et al., 1997). First, we included all models satisfying the following criteria: VIF<5 (to avoid multicollinearity issues); and adjusted R-square>0.2 (adjusted R-square>0.1 for “any treatment” ensemble). Next, we excluded all models receiving less support from the data than their simpler submodels. More complex models (using *n* features) which are less accurate than other any of the less complex models trained on the same subset of features (using any combination of *n-1* features) were excluded. This procedure penalizes more complex models if their accuracy is not superior to simpler models. The pruned ensembles were then sorted by increasing RMSE (decreasing accuracy) and the aggregated output of each ensemble was calculated as the cumulative Bayesian average of individual models for each subject, using the inverse model RMSE as weights (Raftery et al., 1997).

We applied a Bayesian approach for the analysis of coefficients in the pruned ensembles. First, we estimated the prior inclusion probability for an individual feature as the proportion of models including a specific feature relative to the total number of models possible to train under the constraint of maximum 6 features/model to 0.2035 (see Supplementary Material). The posterior inclusion probability (PIP) for each feature was calculated as the frequency of occurrence in the pruned ensembles, and was used for defining levels of evidence strength as follows: PIP>0.2035 identifies features with frequency of occurrence increased as compared to initial probability (enriched), and indicates medium to strong evidence of correlation; in contrast, PIP < prior inclusion probability denotes features depleted after pruning, and indicates weak evidence of correlation. The effect size for individual features was estimated as the average of standardized coefficients across all models in the pruned ensemble. Lastly, we calculated the coefficient of variation (CV=standard deviation/average) for standardized coefficients to estimate the stability of individual features (context-independence).

## Results

### Individual features are differentially affected by MDD treatments

We first evaluated the differences between healthy controls and MDD subjects at population level. Given the differences in inclusion criteria between the ketamine study (insufficient response to SSRI (Tiger et al., 2020)) and the iCBT trial, we evaluated the potential selection bias between the two populations before pooling the data. Sparse between-group differences surviving FDR correction (variability of circadian profile and scaling exponent for short intervals; see Supplementary Fig. S4) did not justify the stratification of MDD patients before treatment into distinct populations. We did not find significant differences between pooled MDD patients before treatment and healthy controls (Fig. 2A). The analysis of the effects of MDD treatments on activity patterns, independently from the magnitude of response, highlighted significant effects on a wide range of features, with distinct, non-overlapping signatures across treatments (Fig. 2A). This approach assumed that MDD treatments, including non-pharmacological intervention (iCBT) and antidepressant drugs (escitalopram, ketamine) may impact circadian regulation of activity independent of antidepressant effects (reviewed in (Kiehn et al., 2019; Silva et al., 2021)). The effects can be summarized as follows: the variability of circadian profile of activity increase in subjects undergoing iCBT. Under escitalopram treatment, the patterns of activity are less variable within the day (lower scaling exponents for short intervals, decreased variability in IV5 and scaling exponent for short intervals), more consistent across days (stronger circadian entrainment, *i.e.*, stable alignment to the 24h light-dark cycle: higher RA and IS30), and the subjects display a higher propensity to sustain activity as compared to untreated MDD subjects. Under ketamine treatment, the patterns of activity are more fragmented (increased IV5), less complex (decreased scaling exponents), and more variable across days (increased circadian profile variability) as compared to untreated MDD subjects. Lastly, comparisons between subjects in remission (regardless the antidepressive treatment leading to remission) and healthy controls or MDD patients before treatment yielded no significant differences for any single feature (Fig. 2A).

**Fig. 2.**
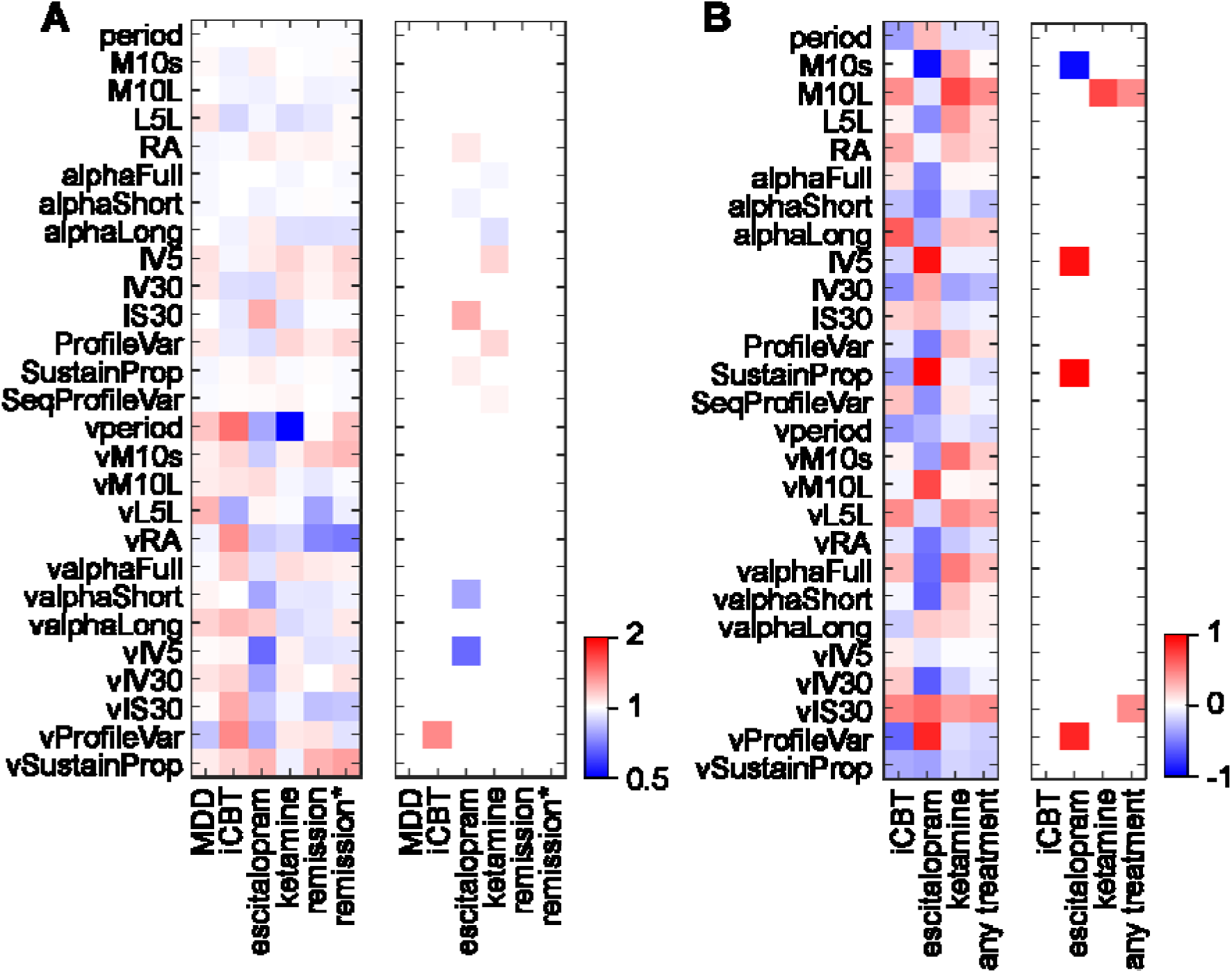
Changes in patterns of activity associated with ongoing antidepressive treatment. (**A**) Between-group differences in individual features. Each treatment group is compared against MDD subjects before treatment as reference. MDD subjects before treatment are compared against healthy controls. The heatmap in the left panel displays the ratios between each group average and their respective reference group average. Significant differences surviving FDR correction are displayed in the right panel. Note the distinct signatures of individual treatments. Subjects in remission (independent of antidepressant treatment) display no significant differences from MDD subjects before treatment (remission) or from healthy controls (remission*). (**B**) Correlations between changes in individual features and the magnitude of response to treatment. Daily variability of IS (vIS30) and timing of circadian peak (M10L) appear to correlate with the response to treatment independently from actual antidepressive treatment.

Next, we evaluated the correlations between the magnitude of response to treatment and longitudinal changes in individual features of activity (Fig. 2B). In subjects treated by iCBT, we did not find significant correlations between the magnitude of response to treatment and changes in individual features, while drug treatments displayed sparse significant correlations with individual features. In subjects treated with escitalopram, better response correlated with increase in circadian peak of activity, decrease in fragmentation of activity, and decrease in propensity to sustain activity. In subjects treated with ketamine infusions, better response to treatment correlated with a shift in location of circadian peak of activity towards earlier occurrence. When all treatments were pooled, better response to treatment was associated with a shift in location of circadian peak of activity towards earlier occurrence, and a stabilization of circadian rhythms (illustrated by decreased daily variability of IS30).

### Distinct signatures in correlations between magnitude of response and individual features for different MDD treatments

We asked whether features of activity patterns during the depressive episode (before initiating any antidepressive treatment) correlate with the magnitude of response to treatment, and whether the correlations are treatment-specific. When each antidepressive treatment was analyzed separately, we found distinct patterns for iCBT, escitalopram, and ketamine (Fig. 3A). For iCBT, better response was associated with higher age, steeper propensity to sustain activity, higher variability in location of circadian trough, and weaker circadian entrainment (lower IS30 and higher vIS30). For escitalopram we did not find correlation surviving correction for FDR, but we observed a trend in correlation between response to treatment and lower variability in fragmentation of activity patterns. Better response to ketamine was associated with increased variability in magnitude of circadian peak of activity. When all cohorts were pooled, better response to treatment was associated with more stable patterns of activity before treatment, as illustrated by decreased variability in location of circadian peak as well as in scaling exponent (Fig. 3A).

**Fig. 3.**
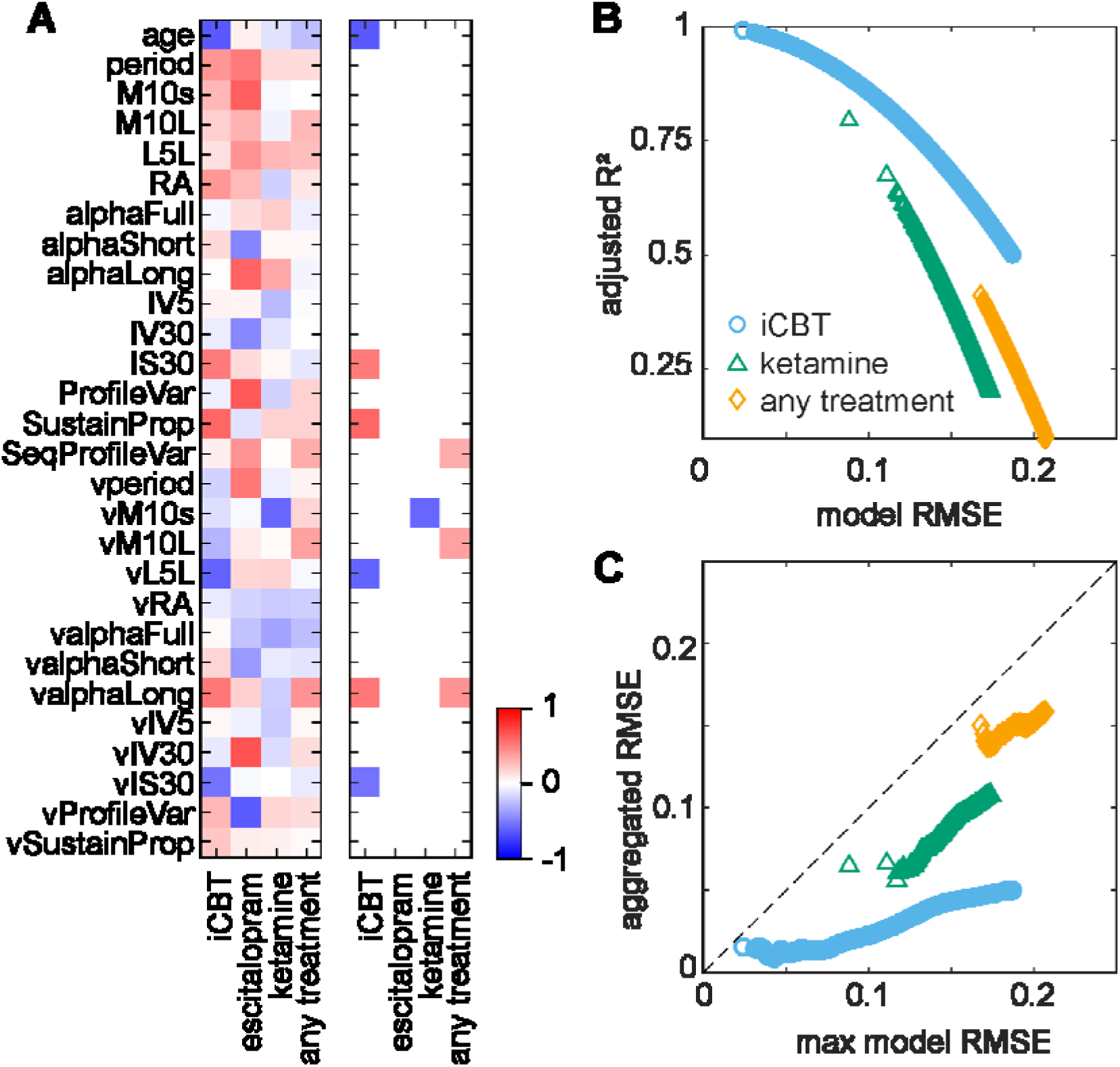
Correlations between activity features before treatment and the response to different antidepressant treatments. (**A**) Correlations with magnitude of response to treatment. Left panel: heatmap of raw correlation coefficients. Right panel: correlation coefficients surviving FDR correction. Note that response to treatment regardless of the nature of antidepressant treatment does not correlate with any individual feature, and that individual treatments have distinct signatures in terms of individual features correlating with the magnitude of the response. (**B**) Parallel training of ensembles of multiple regression models for response to treatment. Illustration of evaluation of performance for individual models. (**C**) Evaluation of performance of pruned ensembles. Aggregated output is calculated as cumulative average of individual model output, after sorting the ensembles by decreasing accuracy (increasing RMSE).

We next asked whether the response to treatment can be modeled by linear combinations of features. To this end, we trained 3 independent ensembles of multiple linear regression models to fit the response to treatment for iCBT and ketamine separately, and used the pooled dataset for fitting the response to treatment irrespective of treatment group (“any treatment”). The pruned ensembles consisted of 41768, 18178, and 23223 models for iCBT, ketamine, and any treatment, respectively (Fig. 3B). To analyze the performance of each ensemble, we sorted the models by decreasing accuracy (increasing RMSE), then calculated the accuracy of ensemble fitting (aggregated output) in a cumulative fashion (Fig. 3C). The aggregated accuracy of all ensembles outperformed independent models, and the accuracy changed with increasing the number of models included in a non-monotonic manner: the initial improvement in accuracy was followed by gradual degradation with different rates for each ensemble (Fig. 3C). Notably, the ensemble fitting the response to any treatment had an accuracy considerably lower than the ensembles fitting the response to either iCBT or ketamine. This is illustrated by the range of variation for aggregated RMSE (minimum: 0.1377 for any treatment, vs. 0.0085 for iCBT and 0.0572 for ketamine; maximum: 0.1588 for any treatment, vs. 0.0496 for iCBT and 0.1092 for ketamine; Fig. 3C). For a MADRS score of 30 before treatment, this translates into an expected error in prediction of MADRS score after treatment between 0.25 and 1.5 for iCBT; 1.7 and 3.3 for ketamine; and 4.1 and 4.8 for any treatment.

To investigate the biological relevance of modeling the response to treatment, we performed a Bayesian analysis of coefficients. The contribution of individual features in ensembles presumably changes depending on the context (*i.e.*, all other features included in the model), and the importance of variables in the model can be assessed using several approaches (Grömping, 2015). We described the contribution of individual features using the following parameters: PIP (*i.e.*, frequency of inclusion in the pruned ensemble, to identify the most relevant features); mean standardized coefficient (to estimate effect magnitude); and coefficient of variance (to estimate the context-dependence for each coefficient) (Fig. 4A; see also Supplementary Fig. S5). These three measurements are not necessarily orthogonal, but their joint analysis highlights the individual contribution of most relevant features. For each ensemble we found enriched features (PIP>0.2035) also had low variability, with limited overlap across ensembles (Fig. 4B). The mean standardized coefficients for individual features varied considerably across ensembles, and filtering the feature space by CV or PIP yielded distinct signatures across ensembles (Fig. 4B; see also Supplementary Fig. S6). Further analyses focused on subsets of enriched features for each ensemble (Fig. 4B and Supplementary Fig. S6). The pattern of correlations between individual features before treatment and the response to treatment can be summarized as follows: better response to iCBT correlated with higher age, weaker circadian entrainment (low IS30, high vM10s, vL5L, vIS30, variability of circadian profile), earlier circadian peak of activity (M10L), and steeper propensity to sustain activity; better response to ketamine correlated with higher age, stronger circadian entrainment (high RA, vM10s, but lower vL5L), higher fragmentation of activity patterns (high IV5), and steeper propensity to sustain activity; and better response to any treatment correlated with higher age, more robust circadian rhythms (high RA), lower and more variable circadian peak of activity (low M10s, high vM10s), and steeper propensity to sustain activity.

**Fig. 4.**
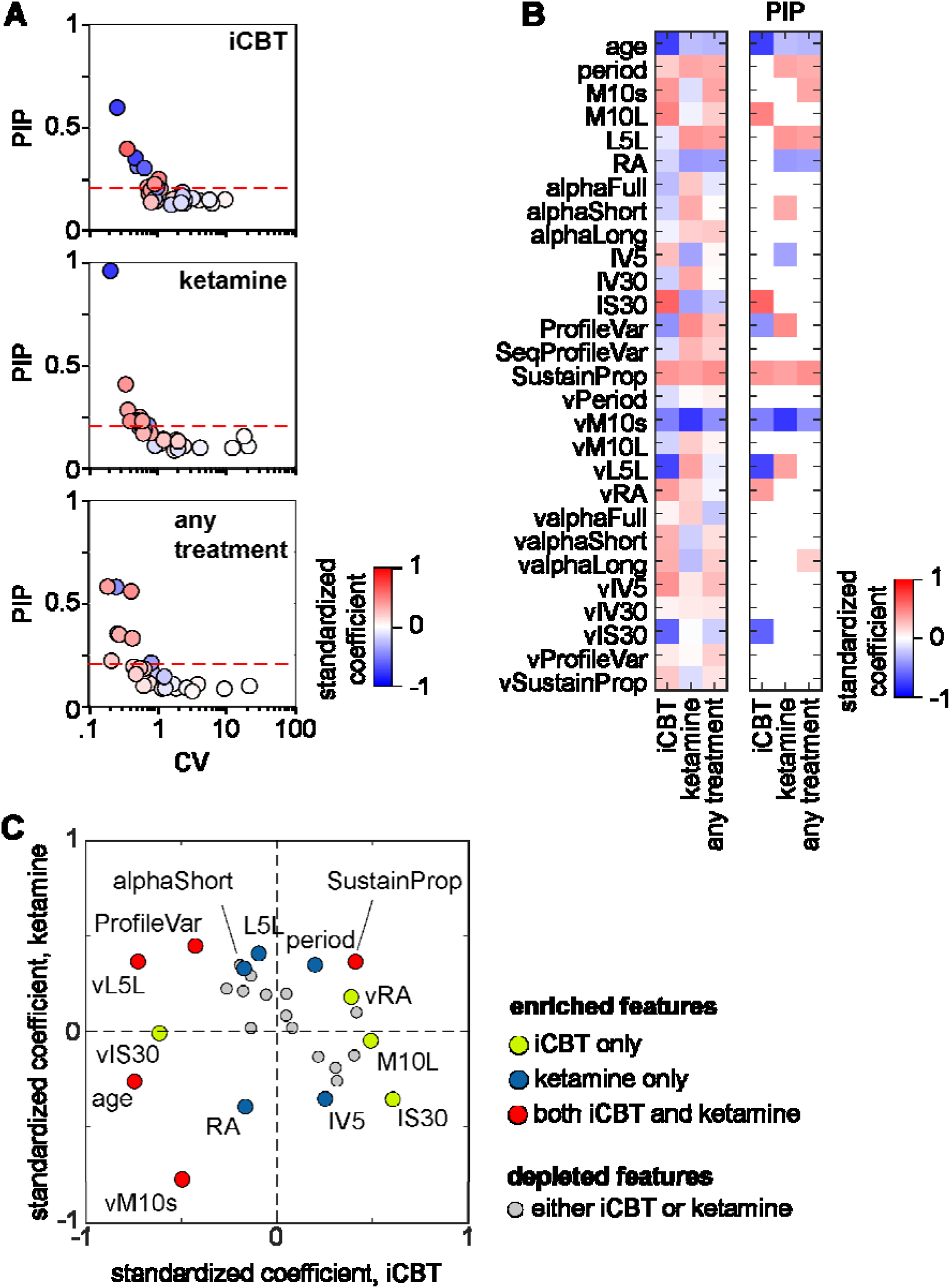
Bayesian analysis of coefficients. (**A**) Analysis of occurrence and variability of individual features in each ensemble. Features mapping above the threshold for PIP (set to prior inclusion probability, 0.2035) are defined as “enriched” since after pruning they are encountered more often than estimated. Features mapping in the top-left region have stable coefficients and have medium to strong evidence of correlation with the magnitude of response to treatment. (**B**) Average standardized coefficients for all ensembles to illustrate the differences in ensemble composition. The heatmap in the first panel shows all average standardized coefficients. The second and third panel display only the coefficients with PIP > 0.2035 (above threshold in (A)). Note the distinct, non-overlapping signatures for each ensemble. (**C**) Direct comparison between effect sizes of individual features in ensembles modeling the response to iCBT and ketamine. Enriched features highlighted and color-coded based on differential enrichment in either ensemble. Features mapping in top-right and bottom-left quadrants indicate similar effects, while features mapping in top-left and bottom-right quadrants indicate opposite effects between ensembles.

To further explore the differences between the ensembles modeling the response to iCBT or ketamine we used a scatterplot to map the contributions of individual features as measured by the standardized coefficients (Fig. 4C). The mapping of enriched features across the quadrants highlighted the distinct contributions of relevant features to modeling the response to treatment: differentially enriched features had larger effects in the ensemble they were enriched in. Out of the 5 features found to be enriched in both ensembles, variability in location of circadian trough and variability in circadian profile had similar effects; while age, variability in circadian peak amplitude, and propensity to sustain activity had opposite effects.

## Discussion

In this study we visualize the impact different MDD treatment alternatives have on circadian patterns of activity. This was possible thanks to the development of a data analysis pipeline, including pre-processing, feature extraction and feature selection for actigraphy recordings as objective measurement. We identified only sparse correlations between individual features measured during a depressive episode and the response to specific MDD treatments, which yielded distinct signatures for each treatment alternative. When the response to treatment was modeled by combinations of features, the Bayesian analysis of coefficients highlighted differences and similarities between contributions of specific features to modelling the response to iCBT or ketamine.

We did not find differences between MDD subjects and healthy controls for individual features, neither before treatment, nor in remission after treatment. MDD is a disease with largely unknown etiology and heterogenous manifestations (according to diagnostic criteria listed in DSM-5^36^, 1497 possible combinations of symptoms can lead to a an MDD diagnosis)(Østergaard et al., 2011), and the diversity within the MDD patient population can confuse comparisons against healthy controls. A large population-based study found that circadian rhythm features (relative amplitude of activity intensity or skin temperature) do not display sex-dependent differences, and have small effect on predicting the diagnosis for mood disorders (Brooks et al., 2021). Similarly, we did not find sex-related differences between groups, nor did we find significant differences between MDD subjects and healthy controls. The ketamine cohort may be assumed fundamentally different from the iCBT and escitalopram cohorts since it was restricted to MDD patients with resistance to SSRI treatment (Tiger et al., 2020). However, we found only sparse differences among cohorts before treatment, which could be related to differences between subgroups being obscured by the overall heterogeneity of the MDD population. In addition, the fact that the healthy controls were matching only the MDD patients in the iCBT cohort, stratification by sex or cohort would not be meaningful when comparing MDD patients against healthy controls.

The analysis of effects of treatment on circadian patterns of activity yielded treatment-specific signatures: iCBT increases variability of circadian profile; escitalopram strengthens circadian entrainment; and ketamine weakens circadian entrainment. For iCBT as non-pharmacological intervention, the patient is supposed to gradually change behavior in a voluntary manner (Weitz et al., 2015). MDD patients undergoing CBT display increased serotonin transporter availability in a composite region including parts of the limbic system (amygdala, cingulate cortex, hippocampus), basal ganglia (caudate and putamen), insular cortex, and thalamus (Svensson et al., 2021), as well as reduced blood concentrations of purine metabolites and increased concentration of branched aminoacids (Bhattacharyya et al., 2024). Alterations in activity patterns can be expected, but there are no definite molecular mechanisms. Circadian entrainment is dependent on the central clock, located in the suprachiasmatic nucleus (SCN) in the anterior hypothalamus, and most neurotransmitter systems on which antidepressants act have been shown to impact circadian rhythms (recently reviewed in (Daut and Fonken, 2019; Lee et al., 2022; Sato et al., 2022)). The effects we observed are in agreement with theoretical pharmacological mechanisms. Serotonin tonically inhibits the direct input from retinal ganglionic neurons, and blocking serotonin reuptake weakens photic entrainment of the SCN (reviewed in (Ciarleglio et al., 2011)). Therefore, blocking photic input to the SCN would simplify the internal clock function, which can explain the strengthening of circadian entrainment we observed in MDD subjects treated with escitalopram. Ketamine has been shown to directly interfere with clock gene transcriptional activity in primary fibroblasts (Bellet et al., 2011), and appears to have widespread suppressive effects on circadian clock machinery (Sato et al., 2022). When applied to the SCN function, two main alterations are expected in patients treated with ketamine: (1) weaker circadian entrainment of activity; and (2) disorganized patterns of activity (see (Hu et al., 2007)). Remarkably, both effects were observed in our cohort. While the pharmacological effects appear consistent at group level, the response ranges from negligible to remission for each treatment. Therefore, we looked for within-patient changes which correlate with “good response”. As expected, the response to specific treatments correlate with changes in different features, but when all cohorts were pooled together, changes in two specific features correlate with the magnitude of response (timing of circadian peak of activity, and variability in circadian entrainment) suggest that stronger circadian entrainment of activity correlates with better response to MDD treatment. In addition, we found that better response correlates with a shift towards earlier occurrence of circadian peak, in agreement with previous reports (Duncan et al., 2017). This points to circadian entrainment of activity being a relevant endpoint for evaluation of response to treatment in MDD subjects, regardless of the antidepressive treatment followed.

We have shown earlier that patterns of activity can be reliably used for modeling symptom severity before treatment (Spulber et al., 2022), and here we hypothesized that the response to treatment, estimated as relative change from baseline, can be modeled using features of circadian patterns of activity during the depressive episode (before treatment initiation). This approach has two potential pitfalls: (1) it does not account for variations in individual items in the MADRS scale; and (2) it ignores all intermediate states between before and after treatment. Regarding the former, it has been shown that the underlying variability is rather limited (Catarino et al., 2022; Rabinowitz et al., 2019; Rabinowitz and Rabinowitz, 2022; Simmonds-Buckley et al., 2021), which makes the total score a robust estimator of current state. We then assumed that symptom severity was reasonably stable (plateau) around the time of recording. The dynamics of response to treatment varies considerably among MDD treatments, but it has been shown to be approximated by an exponential decay (Taliaz et al., 2021). For iCBT, the response appears linear (see (Svensson et al., 2021)), and the response is most reliably evaluated at the end of the course (10 weeks). The response to SSRIs has been described as either fast (stable response is reached within 2 weeks after treatment onset), or slow (symptom severity decreases slowly after an initial plateau), but it is either way stable after 6 weeks of treatment (Fiori et al., 2021). In contrast, a rapid decrease in symptom severity can be documented within hours after a single dose of ketamine administration, followed by a stable response lasting a couple of days (Berman et al., 2000; Sato et al., 2022). We believe that the investigation of correlations between patterns of activity, as expression of current status, and the response to treatment, as change driven by therapeutic intervention, is a meaningful approach, provided the interval between measurements is appropriate for the actual treatment (*i.e.*, 10-12 weeks for iCBT; ∼6 weeks for SSRIs; and ∼2 weeks for ketamine). We have modeled the response to treatment independently from timeline of transition between plateaus using parallel training of linear regression models, then used Bayesian model averaging to account for model uncertainty and increase the generalizability of the findings (Astuti et al., 2014; Raftery et al., 1997). The added value of Bayesian analysis of coefficients is to support the high-level interpretation of correlations between patterns of activity and response to treatment: the standardized coefficients for the subset of enriched features can be translated into behavioral phenotyping of MDD patients who would benefit most from a specific MDD treatment. MDD subjects with weaker circadian entrainment, would have better response to iCBT. However, MDD patients undergoing iCBT appear to increase circadian profile variability, which can be interpreted as weakening of circadian entrainment. Similarly, MDD subjects with fragmented daytime activity would have better response to ketamine, while ketamine treatment increases fragmentation of activity. Taken together, our data indicate that positive effects of specific antidepressive treatment are not necessarily associated with measured changes towards “normalization” of activity patterns (*i.e.*, reverse the deviations from healthy controls). Our data suggest potential prospective applications for predicting the response to specific MDD treatments. However, the generalizability of our findings needs to be tested prospectively on independent datasets.

In conclusion, we identified distinct signatures in the correlations between patterns of activity and response to specific MDD treatments. We further found that the stabilization of circadian entrainment correlates with the magnitude of response independently from the antidepressive treatment followed. Our data suggests that the analysis of circadian patterns of activity may potentially be used for predicting the response to treatment in MDD patients, but larger confirmatory studies are required to support clinical applications.

## Data Availability

All data produced in the present study are available upon reasonable request to the authors

## Acknowledgments

The authors would like to thank Prof. Matteo Bottai (Division of Biostatistics, Karolinska Institutet) for inspiring discussions and suggestions for planning the analyses. Hardware and support for actigraphy data collection was provided by the Ass. Prof. Viktor Kaldo (Department of Psychology, Karolinska Institutet) and Philips (Philips Healthcare, Sleep and Respiratory Care, Murrysville, PA, USA).

## Author contributions

Conceptualization SS, FE, SC, JL. Data curation SS, MG, MT, JL. Formal Analysis SS, FE. Funding acquisition SC, MT, JL. Investigation SS, FE, MG, MT, JL. Methodology SS, FE, MT, JL. Project administration MT, SC, JL. Resources SC, MT, JL. Software SS. Supervision SS, SC, MT, JL. Validation SS, SC, MT, JL. Visualization SS, FE. Writing – original draft SS, FE. Writing – review & editing SS, FE, SC, MG, MT, JL.

## Funding

Funding to support the work was provided by Swedish Society of Medicine, Fredrik and Ingrid Thuring Foundation, Region Stockholm (ALF project 20190429, and clinical research appointment), The Swedish Brain Foundation (Hjärnfonden), and The Söderström König Foundation (SLS-746501) (MT); Swedish Research Council (2013-09304), Region Stockholm (ALF project 20170192 and higher clinical research appointment) (JL); Swedish Research Council (2019-01191), The Swedish Brain Foundation (Hjärnfonden, FO2016-0116), and Torsten Söderberg Foundation (M59/16) (SC); and Karolinska Institutet research grants (MT, SC, JL). The funding agencies and sponsors did not have any influence on the conceptualization, design, data collection, analyses, decision to publish, or preparation of the manuscript.

## Competing interests

The research leading up to the present work generated US Patent No. 10,731,216 – Methods and compositions for biomarkers of depression and pharmacoresponse (SC and SS inventors). The patent is owned by NorthernLight Diagnostics AB (SC and SS co-founders), a company developing decision support tools for mental health care. The company did not provide funding and was not involved in the present study. The other authors declare no competing interests.

## Notes

### Funding Statement

Funding to support the work was provided by The Soederstroem Koenig Foundation (SLS-746501) (MT), Swedish Research Council (2013-09304) (JL); Region Stockholm (ALF project 20170192 and higher clinical research appointment) (JL); Swedish Research Council (2019-01191), The Swedish Brain Foundation (FO2016-0116), Torsten Soederberg Foundation (M59/16) (SC); and Karolinska Institutet research grants (MT, SC, JL). The funding agencies and sponsors did not have any influence on the conceptualization, design, data collection, analyses, decision to publish, or preparation of the manuscript.

### Author Declarations

The studies received ethical approval from Swedish Regional Ethics Committee, Dnr. 2014/452-31, 2015/1177-32, 2017/799-31

### Summary of Updates

revised discussion section for clarity and brevity

