## Supplementary Information for "Correlations between patterns of activity and the response to treatment yield distinct signatures for different antidepressive treatments"

**Supplementary Figures and Tables**

Supplementary Table 1: Group re-allocation and descriptive statistics of data available.

Supplementary Table 2: Complete list of features extracted (separate file).

Supplementary Table 3: Overview of features used for training models.

Supplementary Fig S1: Illustration of the need for range normalization and the effect thereof.

Supplementary Fig S2: Illustration of feature extraction procedure and preliminary analyses before feature selection.

Supplementary Fig S3: Correlations between features before and after feature selection.

Supplementary Fig S4: Differences between MDD subjects included in iCBT and ketamine studies.

Supplementary Fig S5: Synoptic view of the ensembles modeling the response to treatment.

Supplementary Fig S6: Most relevant features modeling the response to treatment.

**Supplementary Material**

Calculation of prior inclusion probability for a specific feature $X_{i}$

$\Pr\left( X_{i} \right)=\frac{N models including X_{i}}{N models possible}$ (1)

For a given complexity level, the total number of models possible to train is equal to the binomial coefficient $\binom{n}{k}$, and the number of models including a specific feature is the binomial coefficient $\binom{n-1}{k-1}$, where n = total number of features available, and k = model complexity (number of features). Therefore equation (1) can be rewritten as follows:

$$\Pr\left( X_{i} \right)=\frac{\sum_{k=0}^{5} \binom{n-1}{k}}{\sum_{k=0}^{6} \binom{n}{k}}$$

Solving numerically for n=28 yields $\Pr\left( X_{i} \right)=\frac{101 584}{499 178}=0.2035$. The prior inclusion probability of 0.2035 applies to all features, because we trained models for all possible combinations of up to 6 features/model. After pruning the model ensembles, enriched features must have PIP>0.2035, while depleted features have PIP<0.2035.

**Supplementary Table 1.** Group re-allocation and descriptive statistics of data available.

| Group | N subjects | N days | days/ M | days/ F |
| --- | --- | --- | --- | --- |
| HC | 19 (14) | 232 (173) | 11.8 | 12.4 |
| MDD (pre)^1^ | 35 (20) | 277 (172) | 6.2 | 8.6 |
| iCBT (post) | 11 (9) | 108 (96) | 6 | 10.7 |
| Escitalopram (post) | 8 (2) | 62 (8) | 10.3 | 4 |
| Ketamine (post) | 15 (6) | 141 (72) | 7.7 | 12 |

Numbers in parentheses indicate the number of days actigraphy recorded in female subjects. See also Fig 1 for details on timing of recording relative to treatment administration.

^1^ – includes all recordings before treatment from all treatment groups

**Supplementary Table 3**

Overview of features used for training models.

| # | feature ID | # | feature ID |
| --- | --- | --- | --- |
| demographic features | | | |
| 1 | age | | |
| calculated on full-length recording | | RMS deviation from average | |
| 2 | period | 16 | vperiod |
| 3 | M10s | 17 | vM10s |
| 4 | M10L | 18 | vM10L |
| 5 | L5L | 19 | vL5L |
| 6 | RA | 20 | vRA |
| 7 | alphaFull | 21 | valphaFull |
| 8 | alphaShort | 22 | valphaShort |
| 9 | alphaLong | 23 | valphaLong |
| 10 | IV5 | 24 | vIV5 |
| 11 | IV30 | 25 | vIV30 |
| 12 | IS30 | 26 | vIS30 |
| 13 | ProfileVar | 27 | vProfileVar |
| 14 | SeqProfileVar |  |  |
| 15 | SustainProp | 28 | vSustainProp |

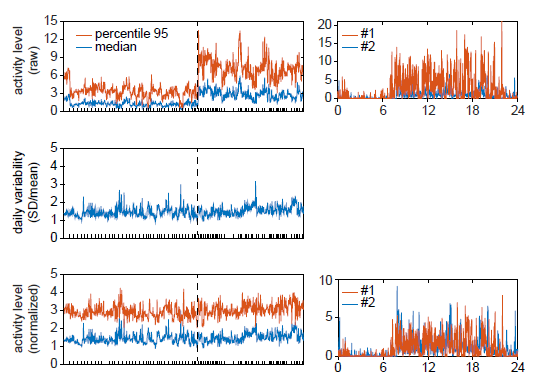
**Supplementary Fig S1: Illustration of the need for range normalization and the effect of normalization.** Top panel: median and 95^th^ percentile of raw activity levels for each day recorded. Tickmarks on horizontal axis show delimitation between individual subjects. Vertical dashed line separates the recordings acquired using different actigraphy devices. Top left panel: illustration of differences between activity counts reported by the 2 recording devices. Middle panel: illustration of within-day variability (coefficient of variation) justifying range normalization. Bottom panels: median and 95^th^ percentile of activity recorded in each day independently after normalization to average. Note the range-match across devices in the bottom right panel.

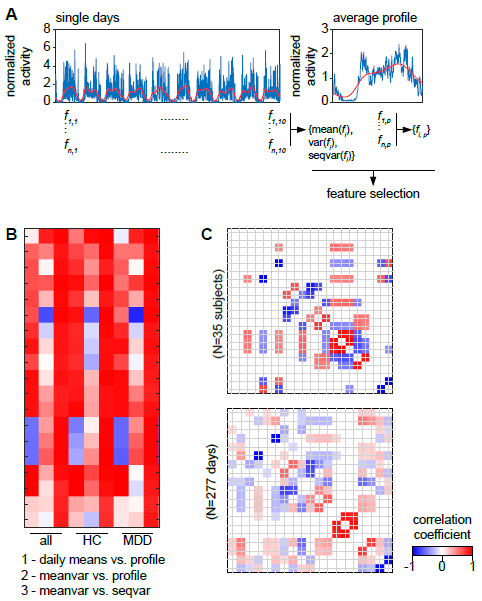
**Supplementary Fig S2: Illustration of feature extraction procedure and preliminary analyses before feature selection.** (A) The same feature set (see Supplementary Table 1 for details) was calculated for each day separately, and for the circadian profile calculated as average of daily profiles. Daily features were further summarized as means, root mean square deviation from average (var), and root mean square sequential differences (seqvar). (B) correlations between the 4 sets of features for all cohorts (all), healthy controls (HC), and MDD patients before treatment (MDD). The purpose of this plot is to identify highly correlated feature sets (i.e., which provide similar information). Note the virtually perfect correlation between average and sequential variations. Note also daily means and profile features are highly correlated, particularly in the MDD group. The decision to include feature sets was taken primarily using the MDD group. (C) Partial correlations between features calculated on average circadian profile (top panel) and individual days (bottom panel) – only correlation coefficients surviving FDR correction shown; MDD subjects. Note that most robust correlations match between the two datasets.

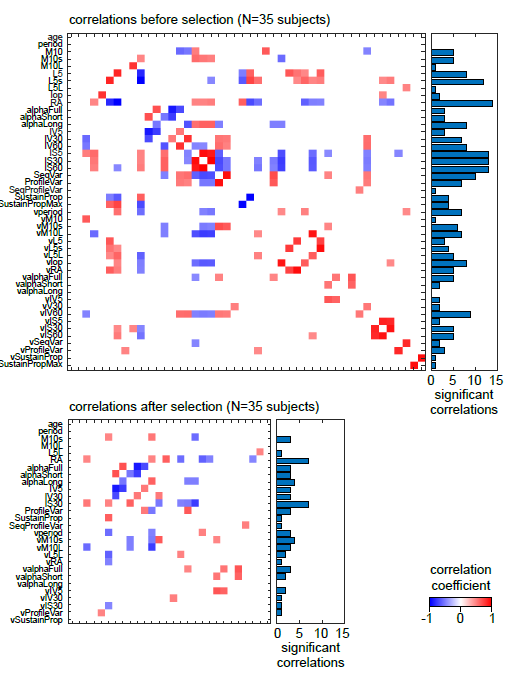
**Supplementary Fig S3: Correlations between features before and after feature selection.** Some features are conceptually related and form highly correlated clusters, visualized as clusters of points along the main diagonal before feature selection (top panel). After selection (bottom panel), the number of significant correlations dropped to below 5 for most features.

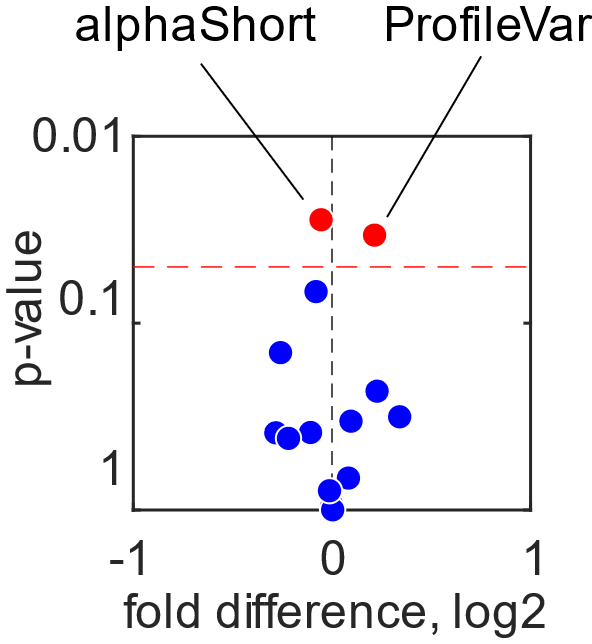

**Supplementary Fig S4: Differences between MDD patients included in the iCBT and ketamine studies.** Volcano plot depicting between-group differences at individual feature level between MDD patients included in the ketamine study as compared to MDD patients included in the iCBT study (used as reference). Significant differences surviving FDR correction found in alphaShort and ProfileVar,

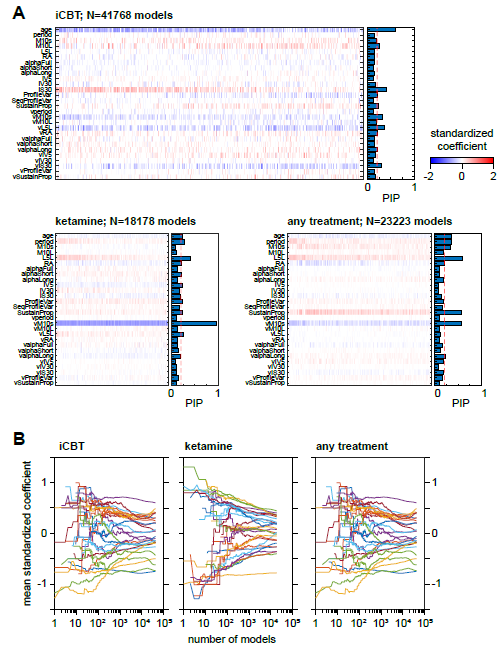
**Supplementary Fig S5: Synoptic view of the ensembles modeling the response to treatment.** (A) Heatmap of standardized coefficients sorted by descending accuracy (increasing RMSE). (B) Cumulative average of standardized coefficients. Note the horizontal axis is logarithmic.

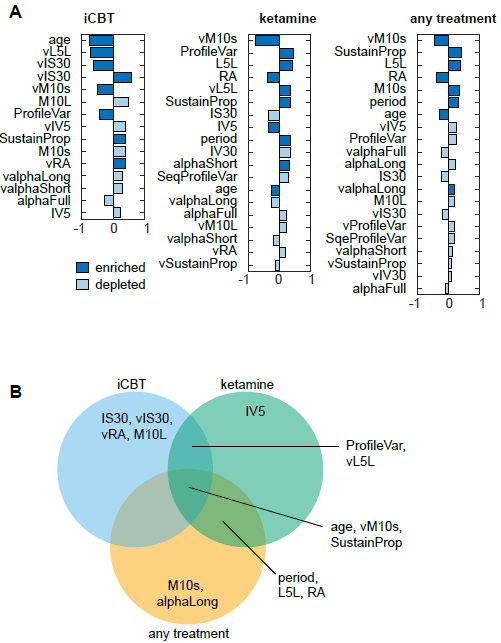
**Supplementary Fig S6: Most relevant features for each ensemble.** Please refer to Fig 4A in the main text for reference. (A) Features with CV>1.5, sorted by absolute effect size. Correlates with weak evidence (*i.e.*, PIP below prior inclusion probability) shown in light blue. Note that age, vM10s, and SustainProp are enriched in all ensembles, and have similar influence (same direction, although different effect size). In contrast, vL5L and ProfileVar are enriched in both iCBT and ketamine ensembles, but have opposite effects. (B) Illustration of enriched features shared among ensembles. Note that the 2 features shared only by iCBT and ketamine have opposite correlations in the two ensembles.
