## Supplementary Table for "Correlations between patterns of activity and the response to treatment yield distinct signatures for different antidepressive treatments"

| Feature | Calculation | Source code | Interpretation |
| --- | --- | --- | --- |
| Circadian period | main peak between 22 and 26 h on Lomb-Scargle periodogram | Adapted from [1] | Circadian period should be ~24h, but varies also depending on the precision of the estimation (oversampling factor) |
| Magnitude of main circadian peak of activity | $M10s=\boldsymbol{max(smoooth} \left( \left\{ \boldsymbol{x}_{\boldsymbol{i}} \right\} \right)\boldsymbol{)}$;  *x_i_*= activity counts in 1 min bins, normalized to daily average; smoothing: sliding gaussian window, 600 samples (10h). | OI; adapted from [2,3] | Magnitude of main circadian peak, normalized to average activity count. |
| Time of occurrence of main circadian peak of activity | M10L=location of M10s (h after midnight) | OI; [3] | Occurs typically in the afternoon. |
| Magnitude of main circadian trough of activity | $L5s=\boldsymbol{min(smoooth} \left( \left\{ \boldsymbol{x}_{\boldsymbol{i}} \right\} \right)\boldsymbol{)}$;  *x_i_*=activity counts in 1 min bins, normalized to daily average; smoothing: sliding 300 samples (5h) gaussian window. | OI; adapted from [2] | Activity during the circadian trough, typically during nighttime rest |
| Time of occurrence of main circadian trough of activity | L5L=Location of L5s (h after midnight) | OI | Occurs typically before 6 a.m. Large deviations may indicate shiftwork. |
| Relative amplitude of circadian rhythms | $RA=\frac{\boldsymbol{M 10}\boldsymbol{s-L}\boldsymbol{5}\boldsymbol{s}}{\boldsymbol{M}\boldsymbol{10}\boldsymbol{s+L}\boldsymbol{5}\boldsymbol{s}}$ | OI; [2,4] | Varies between 0 (constant activity throughout the day) and 1 (no activity whatsoever for at least 5h straight) |
| Scaling exponents | Slope of residual variance (after detrending) against detrending window width. Detrended fluctuation analysis (DFA) on log-equally spaced intervals between 4 and 1448 min (alphaFull); between 4 and 215 min (alphaShort); between 256 and 1448 min (alphaLong). | OI; [5,6] | Evaluation of complexity of patterns of activity. It is a non-linear and non-monotonic scale, where values around 0.5 are typical for random fluctuations, and higher (around 1 and above) indicate complex, self-similar and scale-independent patterns. |
| Intradailiy variability | $IV=\frac{n\sum_{2}^{n} {(b_{i}-b_{i-1})}^{2}}{(n-1)\sum_{1}^{n} {(b_{i}-\bar{b})}^{2}}$  *b_i_*= activity counts binned over 5, 30, or 60 min; *n*=number of samples | OI; [2,7] | Estimates fragmentation of activity. It has been shown that the bin width is relevant for pairwise comparisons between conditions [7], therefore we calculated IV in different bins. |
| Interdailiy stability | $IS=\frac{n\sum_{1}^{p} {(b_{h}-\bar{b})}^{2}}{p\sum_{1}^{n} {(b_{i}-\bar{b})}^{2}}$  *b_i_*= activity counts binned over 5, 30, or 60 min; *b_h_*=average circadian profile; *p*=period (24h); *n*=number of samples | OI; [2,7] | Estimates synchronization with the light-dark cycle, *i.e.*, circadian entrainment. Same reasoning as above to justify the use of multiple bin sizes. |
| Variability of circadian profile – sequential differences | $SeqVar=\frac{\boldsymbol{1}}{\boldsymbol{N-1}}\sum_{\boldsymbol{j=2}}^{\boldsymbol{N}} \sqrt{\frac{\boldsymbol{1}}{\boldsymbol{p}}\sum_{\boldsymbol{i=1}}^{\boldsymbol{p}} {\boldsymbol{(}\boldsymbol{x}_{\boldsymbol{i,j}}\boldsymbol{-}\boldsymbol{x}_{\boldsymbol{i,j-1}}\boldsymbol{)}}^{\boldsymbol{2}}}$  N= number of days; p=period (1440 samples); $\boldsymbol{x}_{\boldsymbol{i,j}}$= activity counts in the *i*-th bin of *j*-th day, normalized to average activity level on *j*-th day. | OI | RMS difference between normalized circadian profiles of consecutive days. |
| Variability of circadian profile – deviation from average profile | $ProfileVar=\frac{\boldsymbol{1}}{\boldsymbol{N}}\sum_{\boldsymbol{j}\boldsymbol{=}\boldsymbol{1}}^{\boldsymbol{N}} \sqrt{\frac{\boldsymbol{1}}{\boldsymbol{p}}\sum_{\boldsymbol{i}\boldsymbol{=}\boldsymbol{1}}^{\boldsymbol{p}} {\boldsymbol{(}\boldsymbol{x}_{\boldsymbol{i}\boldsymbol{,}\boldsymbol{j}}\boldsymbol{-}\bar{\boldsymbol{x}_{\boldsymbol{i}}}\boldsymbol{)}}^{\boldsymbol{2}}}$  N= number of days; p=period (1440 samples); $\boldsymbol{x}_{\boldsymbol{i,j}}$= activity counts in the *i*-th bin of *j*-th day, normalized to average activity level on *j*-th day. | OI | RMS difference between normalized individual days and average normalized circadian profile. |
| Day-to-day variability | $SeqProfileVar=\sqrt{\frac{\sum_{\boldsymbol{j}\boldsymbol{=2}}^{\boldsymbol{N}} \sum_{\boldsymbol{i}\boldsymbol{=1}}^{\boldsymbol{p}} {\boldsymbol{(}\boldsymbol{x}_{\boldsymbol{i}\boldsymbol{,}\boldsymbol{j}}\boldsymbol{-}\boldsymbol{x}_{\boldsymbol{i}\boldsymbol{,}\boldsymbol{j}\boldsymbol{-1}}\boldsymbol{)}}^{\boldsymbol{2}}}{\sum_{\boldsymbol{j}\boldsymbol{=1}}^{\boldsymbol{N}} \sum_{\boldsymbol{i}\boldsymbol{=1}}^{\boldsymbol{p}} {\boldsymbol{(}\boldsymbol{x}_{\boldsymbol{i}\boldsymbol{,}\boldsymbol{j}}\boldsymbol{-}\bar{\boldsymbol{x}_{\boldsymbol{i}}}\boldsymbol{)}}^{\boldsymbol{2}}}}$  N= number of days; p=period (1440 samples); $\boldsymbol{x}_{\boldsymbol{i,j}}$= activity counts in the *i*-th bin of *j*-th day, normalized to average activity level on *j*-th day. | OI | Root ratio between squared sequential differences and the squared deviations from average profile (hence cannot be calculated for individual days). For random samples, SeqProfileVar is approximately $\sqrt{2}$. It decreases if the circadian profile is consistent (*i.e.*, small differences between consecutive days). |
| Propensity to sustain activity | $\boldsymbol{P(x}_{\boldsymbol{i+1}}\boldsymbol{\geq}\boldsymbol{x}_{\boldsymbol{i}}\left\vert\boldsymbol{x}_{\boldsymbol{i}}\boldsymbol{\in}\boldsymbol{b}_{\boldsymbol{j}} \right)\boldsymbol{=m*}\boldsymbol{center(b}_{\boldsymbol{j}}\boldsymbol{)+n}$  *SustainProp=****m***; *SustainPropMax*=***n***  *{b_j_}*= distribution of activity in log-equally spaced bins. | OI | The intercept (***n***) can be interpreted as the likelihood to increase activity from 1 activity count/min, but has little biological relevance given that the distribution is truncated to remove very low activity levels associated with resting/sleep. In addition, the regression line will always approach 0 towards the right tail of the distribution, therefore slope (***m***) and intercept (***n***) are highly correlated. |

**Abbreviations:** OI – own implementation in Matlab™, adapted from published reports or original contribution if no reference provided.
